# Association of contact to small children with mild course of COVID-19

**DOI:** 10.1101/2020.07.20.20157149

**Authors:** Martin Dugas, Inga-Marie Schrempf, Kevin Ochs, Christopher Frömmel, Leonard Greulich, Philipp Neuhaus, Phil-Robin Tepasse, Hartmut H-J Schmidt

## Abstract

It is known that severe COVID-19 cases in small children are rare. If a childhood-related infection would be protective against severe course of COVID-19, it would be expected that adults with intensive and regular contact to small children also may have a mild course of COVID-19 more frequently. To test this hypothesis, a survey among 4,010 recovered COVID-19 patients was conducted in Germany. 1,186 complete answers were collected. 6.9% of these patients reported frequent and regular job-related contact to children below 10 years of age and 23.2% had own small children, which is higher than expected. In the relatively small subgroup with intensive care treatment (n=19), patients without contact to small children were overrepresented. These findings are not well explained by age, gender or BMI distribution of those patients and should be validated in other settings.

## Introduction

The coronavirus disease 2019 (COVID-19) pandemic has affected millions of people worldwide. In contrast to many other infectious diseases, children have diagnostic findings similar to adults, but fewer children seem to have developed severe pneumonia [1]. At present, this phenomenon is not well understood. One of several explanations might be that infections which are frequent in childhood are protective against severe course of COVID-19.

There are recent reports about cross reactivity regarding COVID-19. For instance, it was shown that T cell reactivity to SARS-CoV-2 epitopes is also detected in non-exposed individuals [2]. In principal, cross reactivity might contribute to the heterogeneity of COVID-19 infections: The majority of patients can be treated in an outpatient setting, but some patients require long-term intensive care.

If a childhood-related infection would be protective against severe course of COVID-19, it would be expected that adults with intensive and regular contact to small children also may have a mild course of COVID-19 more frequently, because these adults are more exposed to those childhood-related infections. To test this hypothesis, a survey among recovered COVID-19 patients was conducted.

## Methods

In the context of the Coronaplasma Project (local ethic committee approval: AZ 2020-220-f-S) at the University Hospital of Münster, Germany, an online-survey of 4,010 persons who recovered from a confirmed COVID-19 infection was performed. This cohort volunteered to donate plasma with antibodies against COVID-19. Most of these persons live in the state of North Rhine-Westphalia in Germany. Because only few cases with severe course of disease are contained in this cohort, an additional group of 9 COVID-19 patients requiring intensive care therapy at the University Hospital in Münster were included in this survey.

The main question was: Did You have frequent and regular contact to children below 10 years?

Answer options were: Yes, due to my job (in particular kindergarten, primary school, pediatric practice); Yes, due to my own children; No.

These data were combined with basic demographic data (age, gender), body mass index (BMI) and type of COVID-19 treatment (outpatient only; inpatient; inpatient with intensive care).

## Results

2,112 answers were collected with the online survey (response rate 52.7%). 1,186 records (effective response rate 29.6%) contained all required data items and were further analyzed. Median age of participants was 46 years (range 17 - 77). 56% were females, 44% males. Median body mass index was 24.7. 82 persons (6.9%) reported frequent and regular contact to children below 10 years due to their job. 276 persons (23.2%) answered frequent and regular contact to their own children below 10 years of age. 59 (5.0%) reported inpatient treatment, 19 (1.6%) were admitted to intensive care units (ICU). 18 of 19 (95%) ICU patients were males.

Age in the group with regular contact to children (n=358, “child group“) was lower compared to the group without this type of contact (n=828, “nochild group“): median 41 versus 49 years (p=0.002). BMI was slightly higher in the child group (median 25.2 versus 24.6). The proportion of females was 60% in the child group and 54% in the nochild group. Regarding inpatient treatment there was no difference between both groups: 19 (5.0%) in the child group, 40 (4.8%) in the nochild group. Of note, intensive care treatment was reported by 3 of 19 (16%) inpatients in the child group and by 16 of 40 inpatients (40%) in the nochild group (p=0.056, exact Fisher test, one-sided). Median age of ICU patients in the child group was 55, in the nochild group 59.5 years.

## Discussion

Two findings of this survey are noteworthy: First, 6.9% of a large cohort with predominantly mild course of COVID-19 (no fatalities, only 1.6% with intensive care treatment) reported frequent and regular job-related contact to children below 10 years of age. According to the German national statistics agency, there are approximately 650.000 kindergarten teachers, 230.000 primary school teachers and 15.000 pediatricians for a country with 83 million people [3,4,5], i.e. approximately 1% of the population has frequent job-related contact to small children. Our COVID-19 cohort is not a representative sample of the general population, but the rate of job-related contacts to small children seems to be elevated, which is not well explained by age, gender or BMI distribution of this cohort. Altogether, approximately 30% of our cohort report frequent contact to small children, which is higher than in the general population.

Second, in the relatively small subgroup with intensive care treatment, patients without contact to small children were overrepresented. Again, this is not well explained only by age, gender or BMI distribution for this subgroup.

This survey has important limitations: Our cohort is not a representative sample of the general population. Like in many surveys, the response rate is limited and evaluable data was only available for about 30% of the contacted persons. Data were provided by laypersons and are not validated by medical professionals. And importantly, association is not causation.

What can be concluded from this survey and what are possible next steps? It is known that severe COVID-19 cases in small children are rare. There seems to be an association of contact to small children with mild course of COVID-19 in adults. This finding needs to be validated in other settings. An obvious hypothesis derived from this data is that certain childhood infections might provide partial immunity against COVID-19 and thereby potentially could reduce the need for ICU therapy in adults. To test this hypothesis, laboratory tests for suitable pathogens should be performed, preferably with biomaterial collected before the COVID-19 infection. Candidates for such pathogens include endemic coronaviruses like HCoV-NL63 and -229E, which are commonly found in children. From a public health perspective, specific biomarkers to predict the course of COVID-19 *before* an infection would be highly valuable to identify patients at risk, because even healthy young and middle-aged adults can die from COVID-19. If cross reactivity with relatively harmless pathogens like endemic coronaviruses would be demonstrated, this might even become a vaccination strategy - like vaccination with cow pox helped to eradicate smallpox in the last century.

## Data Availability

Due to data protection regulations,
individual patient data cannot be published for this study.

## References

1. Ludvigsson JF. Systematic review of COVID-19 in children shows milder cases and a better prognosis than adults. Acta Paediatr. 2020;109(6):1088–1095. doi:10.1111/apa.15270

2. Grifoni A, Weiskopf D, Ramirez SI, et al. Targets of T Cell Responses to SARS-CoV-2 Coronavirus in Humans with COVID-19 Disease and Unexposed Individuals. Cell. 2020;181(7):1489-1501.e15. doi:10.1016/j.cell.2020.05.015

3. https://de.statista.com/statistik/daten/studie/1011406/umfrage/fachkraefte-in-der-kinderbetreuung-in-deutschland/ [accessed July 16, 2020]

4. https://de.statista.com/statistik/daten/studie/162263/umfrage/anzahl-der-lehrkraefte-nach-schularten/ [accessed July 16, 2020]

5. https://de.statista.com/statistik/daten/studie/158849/umfrage/aerzte-nach-taetigkeitsbereichen-in-deutschland/ [accessed July 16, 2020]

